# High titers of infectious SARS-CoV-2 in COVID-19 corpses

**DOI:** 10.1101/2022.10.11.22280868

**Authors:** Hisako Saitoh, Yuko Sakai-Tagawa, Sayaka Nagasawa, Suguru Torimitsu, Kazumi Kubota, Yuichiro Hirata, Kiyoko Iwatsuki-Horimoto, Ayumi Motomura, Namiko Ishii, Keisuke Okaba, Kie Horioka, Hiroyuki Abe, Masako Ikemura, Hirofumi Rokutan, Munetoshi Hinata, Akiko Iwasaki, Yoichi Yasunaga, Makoto Nakajima, Rutsuko Yamaguchi, Shigeki Tsuneya, Kei Kira, Susumu Kobayashi, Go Inokuchi, Fumiko Chiba, Yumi Hoshioka, Aika Mori, Isao Yamamoto, Kimiko Nakagawa, Harutaka Katano, Shun Iida, Tadaki Suzuki, Shinji Akitomi, Iwao Hasegawa, Tetsuo Ushiku, Daisuke Yajima, Hirotaro Iwase, Yohsuke Makino, Yoshihiro Kawaoka

**Author notes:** **Corresponding Author:** Hisako Saitoh, Department of Legal Medicine, Graduate School of Medicine, Chiba University, 1-8-1 Inohana, Chuo-ku, Chiba 260-8670, Japan.

## Abstract

**Background:** The prolonged presence of infectious severe acute respiratory syndrome coronavirus (SARS-CoV-2) in deceased coronavirus disease 2019 (COVID-19) patients has been reported. However, infectious virus titers have not been determined. Such information is important for public health, death investigation, and handling corpses.

**Aim:** The aim of this study was to assess the level of SARS-CoV-2 infectivity in COVID-19 corpses.

**Methods:** We collected 11 nasopharyngeal swabs and 19 lung tissue specimens from 11 autopsy cases with COVID-19 in 2021. We then investigated the viral genomic copy number by real-time reverse transcription-polymerase chain reaction and infectious titers by cell culture and virus isolation.

**Results:** Infectious virus was present in 6 of 11 (55%) cases, 4 of 11 (36%) nasopharyngeal swabs, and 9 of 19 (47%) lung specimens. The virus titers ranged from 6.00E + 01 plaque-forming units (PFU)/mL to 2.09E + 06 PFU/g. In all cases in which an infectious virus was found, the time from death to discovery was within 1 day and the longest postmortem interval was 13 days.

**Conclusion:** COVID-19 corpses may have high titers of infectious virus after a long postmortem interval (up to 13 days). Therefore, appropriate infection control measures must be taken when handling corpses.

## Introduction

Coronavirus disease 2019 (COVID-19) has caused more than 6.4 million deaths as of September 2022, and the causative pathogen, severe acute respiratory syndrome coronavirus (SARS-CoV-2), continues to circulate worldwide. When autopsies are performed on bodies that have or are suspected to have been infected with this novel coronavirus, detecting SARS-CoV-2 and evaluating its infectivity is important to better determine the cause of death (Deinhardt-Emmer et al., 2021; Fitzek et al., 2021). During autopsies, it is also important to reduce the risk of infection for personnel involved in the examination (Gabbrielli et al., 2021; Schröder et al., 2021). Knowing the infectivity of viruses that remain in corpses is a key public health issue for pathologists, forensic pathologists, medical examiners, clinical physicians, and nurses in addition to those that handle corpses.

Heinrich et al. (2021) detected subgenomic RNA, which indicates viral replication, in specimens collected from the pharynx of COVID-19 corpses as much as 35.8 hours after death. Plenzig et al. (2021) reported that infectious SARS-CoV-2 was present in the lungs of two corpses with postmortem times of 4 and 17 days, respectively. Zacharias et al. (2021) reported that seven of 11 lung tissue swabs were culture positive, with a postmortem interval (PMI) ranging from 14 to 68 hours. Heinrich et al. (2022) further reported that 20% of 128 SARS-CoV-2 RNA-positive corpses retained infectious virus up to 14 days after death.

These reports describe the presence of subgenomic RNA, demonstrating virus replication in the samples or live virus. However, the amounts of infectious SARS-CoV-2 in corpse tissues have not been determined. Therefore, here we evaluated infectious virus titers of samples taken from COVID-19 corpses autopsied in Japan between January and October 2021. We further examined the relationship between the infectivity of the corpse and the viral RNA load on the autopsy date, the time from COVID-19 diagnosis to death, the time in the mortuary, and the PMI.

## Materials and methods

### Samples

A total of 30 specimens (11 nasopharyngeal swabs and 19 lung tissues) were collected from forensic and pathological autopsies of COVID-19 corpses in Japan between January and October 2021. Three lung tissue specimens from the 11 cases were not tested. Information on the age, sex, body mass index (kg/m^2^), cause of death, place of death, date of death (month), decomposition status at the time of autopsy, days from COVID-19 diagnosis to death, and PMI for each case is shown in Table 1. The date of COVID-19 diagnosis is defined here as the date of the first positive result by polymerase chain reaction (PCR) testing or as the date of death unless diagnosed before death.

**Table 1.** Information for 11 autopsy cases with COVID-19.

| Case No. | Age range<br>(yrs) | Sex | BMI<br>(kg/m<sup>2</sup>) | Cause of death | Place of death | Month of death | Decomposition<br>status | From<br>COVID-19<br>diagnosis<br>to death (days)<br>(A) | From death<br>to discovery<br>(days)<br>(B) | Days in<br>morgue<br>(C) | Postmortem<br>interval<br>(B+C) | From<br>COVID-19<br>diagnosis<br>to autopsy<br>(days)<br>(A+B+C) |
| --- | --- | --- | --- | --- | --- | --- | --- | --- | --- | --- | --- | --- |
| 1 | 71-75 | M | 22.4 | Drowning | Pond<br>(4 days after discharge<br>from hospital) | January | None | 13 | 4 | 7 | 11 | 24 |
| 2 | 76-80 | M | 25.9 | COVID-19 pneumonia,<br>Liver cirrhosis | Hospital | February | None | 3 | 0 | 5 | 5 | 8 |
| 3 | 56-60 | F | 30.5 | COVID-19 pneumonia | Hotel (recuperation<br>without clinical treatment) | March | None | 10 | 1 | 2 | 3 | 13 |
| 4 | 56-60 | M | 28.4 | COVID-19 pneumonia | Hotel (recuperation<br>without clinical treatment) | May | None | 4 | 1 | 4 | 5 | 9 |
| 5 | 86-90 | F | 17.6 | COVID-19 pneumonia | Hospital | August | None | 8 | 0 | 4 | 4 | 12 |
| 6 | 26-30 | M | 38.6 | COVID-19 pneumonia | Home (outpatient) | August | None | 2 | 0 | 8 | 8 | 10 |
| 7 | 36-40 | M | 27.6 | Possible COVID-19<br>pneumonia | Home (outpatient) | August | Mild | 9 | 3 | 3 | 6 | 15 |
| 8 | 61-65 | F | 20.9 | Possible COVID-19<br>pneumonia | Home (outpatient) | August | Severe | 3 | 4 | 7 | 11 | 14 |
| 9 | 71-75 | M | 22.5 | Possible COVID-19<br>pneumonia | Home (no medical<br>examination) | August | Severe | 0 | 24 | 2 | 26 | 26 |
| 10 | 71-75 | M | 27.5 | COVID-19 pneumonia | Home (no medical<br>examination) | September | None | 0 | 1 | 12 | 13 | 13 |
| 11 | 36-40 | M | 25.4 | COVID-19 pneumonia | Home (no medical<br>examination) | October | None | 0 | 1 | 12 | 13 | 13 |

### Sample collection

Nasopharyngeal swabs were collected just before the autopsy. The swabs were placed in culture medium (SUGIYAMA-GEN Co., Ltd. Tokyo, Japan) and kept on ice during the autopsy and then stored at -80 °C when the autopsy was completed. An approximately 1–2 cm^2^ pulmonary tissue sample was dissected with scissors and tweezers that were pre-wiped with 70% ethanol, kept on ice during the autopsy, and stored at -80 °C after autopsy completion. Frozen tissues were homogenized and a clarified supernatant was used for virus isolation.

### RNA extraction

RNA was extracted from a 200-µL nasopharyngeal swab culture medium sample or approximately 10 mg of pulmonary tissue sample by using the Maxwell^®^ RSC Viral Total Nucleic Acid Purification Kit (Promega, Madison, Wisconsin, USA).

### Virus quantification by real-time reverse transcription-polymerase chain reaction (RT-PCR)

Real-time RT-PCR of the N1 and N2 viral genomic regions was performed using 1 µL of the RNA preparation. The probe and primer sequences and reaction conditions were as previously published by Adachi et al. (2020) and Shirato et al. (2019). Endogenous hGAPDH-mRNA was used as an internal control, as previously described by Katano et al. (2011).

### Cell culture and virus isolation

Cell culture and virus isolation were performed according to the method of Matsuyama et al. (2020). VeroE6/TMPRSS2 (JCRB1819) cells were obtained from the National Institutes of Biomedical Innovation, Health and Nutrition, Japan. Cells were maintained in Dulbecco’s modified Eagle’s medium (DMEM) containing 10% fetal calf serum (FCS) and antibiotics at 37 °C with 5% CO_2_.

A 24-well plate containing a VeroE6/TMPRSS2 cell culture monolayer at a low crowding density (70–90% confluence) was prepared. After the medium was discarded, 100 μL of the undiluted sample or 10-fold dilutions of the sample were added to the cells and incubated for 1 h at 37 °C. Then, 0.5 mL of DMEM with 5% FCS and antibiotics was added and incubated at 37 °C for 1 week until a cytopathogenic effect (CPE) was observed.

### Virus titration assay

Confluent Vero E6/TMPRSS2 cells in 12-well plates were infected with 100 µL of undiluted or 10-fold dilutions (10^−1^ to 10^−5^) of the sample. After incubation for 1 h at 37 °C, the cells were washed once and overlaid with 1% agarose solution in DMEM with 5% FCS. The plates were incubated for 3 days and then fixed with 10% neutral buffered formalin. After removal of the agar, the plaques were counted.

### Biosafety statement

All experiments with SARS-CoV-2 viruses were performed in the University of Tokyo’s enhanced biosafety level 3 containment laboratories, which are approved for such use by the Ministry of Agriculture, Forestry, and Fisheries, Japan.

## Results

### Viral genomic copy number and infectious virus titers

Infectious virus was detected in 6 (Cases 2, 3, 4, 6, 10, and 11) of 11 cases and 13 (four nasopharyngeal swabs and nine lung tissue specimens) of 30 specimens (Table 2). Figure 1 shows the relationship between viral genomic copy number on the autopsy date, time (days) from the COVID-19 diagnosis to the autopsy date, and the presence or absence of infectious virus. Infectious virus titers ranged from 6.00 E + 01 plaque-forming units (PFU)/mL to 6.00 E + 03 PFU/mL in nasopharynx swabs and from 3.89 E + 02 PFU/g to 2.09 E + 06 PFU/g in lung tissues. The viral genomic copy number for the specimens ranged from the third power of 10 to the seventh power of 10, except in Case 1. Of the 13 specimens containing infectious virus, the lowest viral genomic copy number (3,840 copies/µL) was found in the left lung tissue of Case 11, which had an infectious titer of 1.27E + 04 PFU/g. Of the 17 specimens containing non-infectious virus, the highest viral genomic copy number (7.23E + 06 copies/µL) was found in the left lung tissue of Case 5.

**Figure 1.**
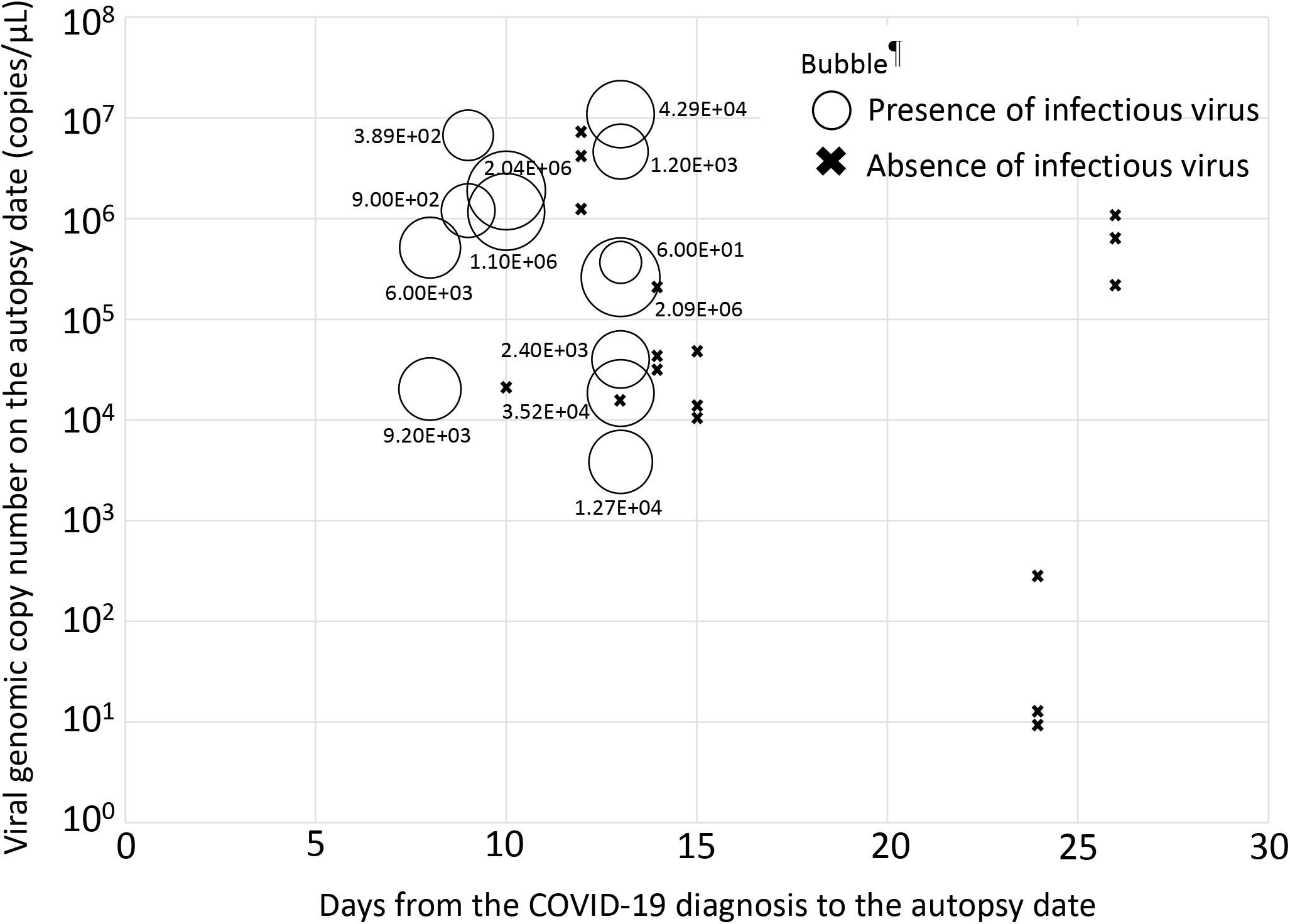
Bubble chart showing the relationship between viral genomic copy number on the autopsy date, days from the COVID-19 diagnosis to the autopsy date, and the presence or absence of infectious virus. ¶The numbers around the bubble indicate the infectious titer and the size of the bubble represents the magnitude of the infectious titer.

**Table 2.**
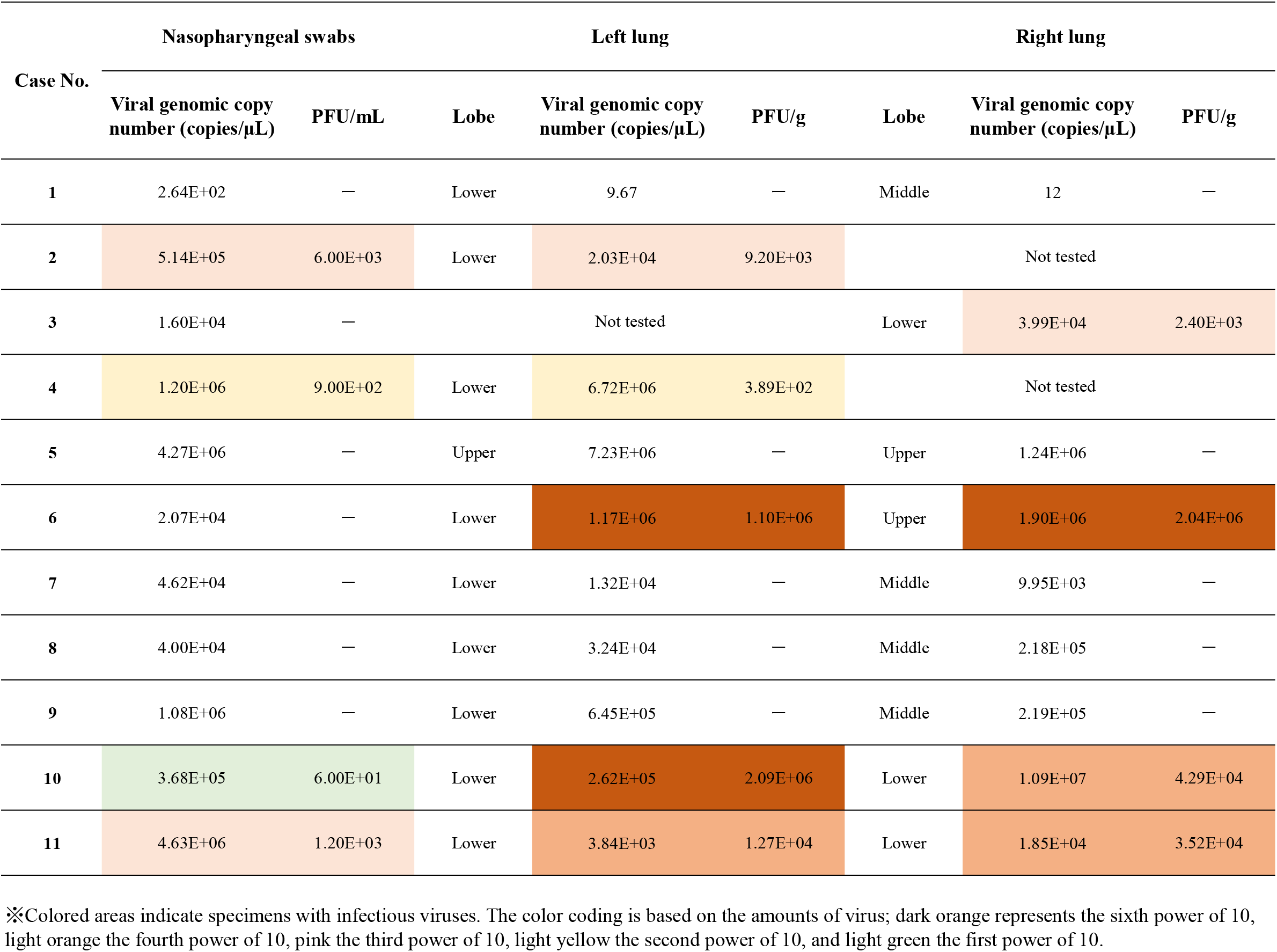
Viral genomic copy number and virus titer.

Of the two hospitalized cases (Cases 2 and 5), infectious virus was found in Case 2, in which the patient had been hospitalized for cirrhosis, contracted COVID-19 in the hospital, and died of severe pneumonia. In Case 5, the patient was hospitalized for 8 days after COVID-19 diagnosis, died despite clinical treatment, and was autopsied 4 days after death. In this case, the viral genomic copy number was in the million-copy range for the nasopharyngeal swab and the left and right lungs, but none of the specimens contained infectious virus.

No infectious virus was detected in Case 1, for which the viral genomic copy number was low, but this body was found in a pond 4 days after discharge when COVID-19 was already cured; the cause of death was drowning.

Of the eight non-hospitalized patients, virus was isolated in five cases (Cases 3, 4, 6, 10, and 11) and no infectious virus was detected in the other three cases (Cases 7, 8, and 9).

In two cases (Cases 3 and 4), the patients had been recuperating in hotel rooms without clinical treatment and were found to contain infectious virus. Among the three cases (Cases 6, 7, and 8) of patients that were recovering at home, the corpse in Case 6 contained infectious virus whereas the other two cases did not. The patient in Case 6 had a body mass index of 38.6 (i.e., was obese) and died at home 2 days after COVID-19 diagnosis. In contrast, the patient in Case 7 died 9 days after diagnosis and was found 3 days after death, and the patient in Case 8 died 3 days after diagnosis and was found 4 days after death in a highly decomposed state.

In the three cases (Cases 9, 10, and 11) of persons found dead at home, the corpses in Cases 10 and 11 were found to contain infectious virus; however, both were in poor health before death and were found to be positive by postmortem PCR testing. The corpse in Case 9 was in a highly decomposed state and infectious virus was not found.

### State of the corpses

Infectious virus was detected in three (Cases 2, 4 and 6) of the four cases (Cases 2, 4, 6, and 8) for which 4 days or less had elapsed from antemortem diagnosis to death.

The two cases (Cases 10 and 11) of three cases (Cases 9, 10 and 11) in which infection was detected by postmortem PCR testing were stored in a refrigerator the day after death; the left and right lungs of both corpses showed high infectious titers. These two cases were not decomposed.

In the two cases (Cases 8 and 9) for which infectious virus was not detected, the corpses had been left at room temperature before discovery and were highly decomposed.

Virus was isolated from six of the seven cases in which the time from death to discovery was within 1 day. In these six cases, the period of refrigeration ranged from 2 to 12 days and the PMI ranged from 3 to 13 days. Case 5 was the only case from which the virus was not isolated; the patient in this case died 8 days after starting COVID-19 treatment.

## Discussion

Here, we determined the amount of virus in COVID-19 corpses and found that infectious virus was present in large amounts (up to 2.09E + 06 PFU/g) in corpse lung tissue and in refrigerated corpses even 13 days after death. Our findings indicate that individuals involved in autopsies or examinations of corpses must consider the risk of infection with SARS-CoV-2 and take measures to protect themselves from infection to reduce that risk. We believe that our findings are crucial for public health globally.

In our limited number of samples, we found that when death occurs because of rapidly worsening COVID-19 symptoms or within a short period after SARS-CoV-2 infection, the probability that infectious virus remains in the corpse is high. If antemortem information is not available, postmortem SARS-CoV-2 PCR testing should be performed. If SARS-CoV-2 PCR-positive corpses are found within a few days after death and they are kept in a cold environment, they should be handled with caution due to likely presence of infectious virus. Autopsies of bodies infected or suspected of being infected with SARS-CoV-2 should be performed in accordance with the postmortem guidance of the World Health Organization (2020) and the Centers for Disease Control and Prevention (2022).

In our study, the titer in lung tissue was higher than that in the nasopharyngeal swab fluid. Therefore, great care should be taken when handling the lungs during autopsy and formalin fixation should be performed immediately. Also, appropriate infection control measures should be implemented during the entire funeral and burial processes to protect workers handling corpses. To this end, ensuring that all those involved in handling corpses are promptly provided with the necessary supplies for infection protection is critical. Furthermore, measures such as prioritizing vaccination in this population of workers should be considered.

## Limitations

The limitation of this study is the small number of COVID-19 corpses examined. In addition, antemortem information was insufficient in some cases.

## Conclusion

To the best of our knowledge, this study is the first to show infectious virus titers in COVID-19 corpses. SARS-CoV-2 remains infectious with cold storage regardless of the PMI for at least 13 days. Therefore, those involved in the autopsies or examinations of corpses must consider the risk of infection with SARS-CoV-2 and take appropriate measures to protect themselves from infection to reduce that risk.

## Data Availability

All data produced in the present work are contained in the manuscript.

## Ethics statement

The study protocol was approved by the Institutional Review Boards of Chiba University Graduate School of Medicine, The University of Tokyo Graduate School of Medicine, and the Institute of Medical Science.

## Author agreement

All authors have seen and approved the final version of the manuscript being submitted. The article is the authors’ original work, has not been previously published, and is not under consideration for publication elsewhere.

## Acknowledgments

We offer our condolences to the families and friends of all the patients whose deaths were attributed to COVID-19. We express our deep thanks to Dr. Rintaro Sawa at the Japan Medical Association Research Institute for advice regarding this project. We thank Dr. Susan Watson, Anahid Pinchis from Edanz (https://jp.edanz.com/ac) for editing a draft of this manuscript.

## Author contributions

Conception and design of the study, HS, SN, TS, SA, TU, HA, MI, DY, HI, YM, and YK; acquisition of data, YST, KIH, HK, SI, Y. Hirata, A. Mori, A. Motomura, HR, MH, AI, YY, MN, RY, S.Tsuneya, K.Kira, SK, GI, FC and Y. Hoshioka; analysis and interpretation data, K. Kubota, S.Torimitsu, NI, KO, KH, IY, KN, and IH.

Drafting the article, HS, SN, SA, K.Kubota, S,Torimitsu, and YM; and revising the article, TS, TU, HA, MI, DY, HI, YST, KIH, HK, SI, Y. Hirata, A.Mori, A. Motomura, HR, MH, AI, YY, MN, RY, S.Tsuneya, K.Kira, SK, GI, FC, Y.Hoshioka, NI, KO, KH, IY, KN, IH, and YK.

All authors approved the final version.

## Graphical abstract

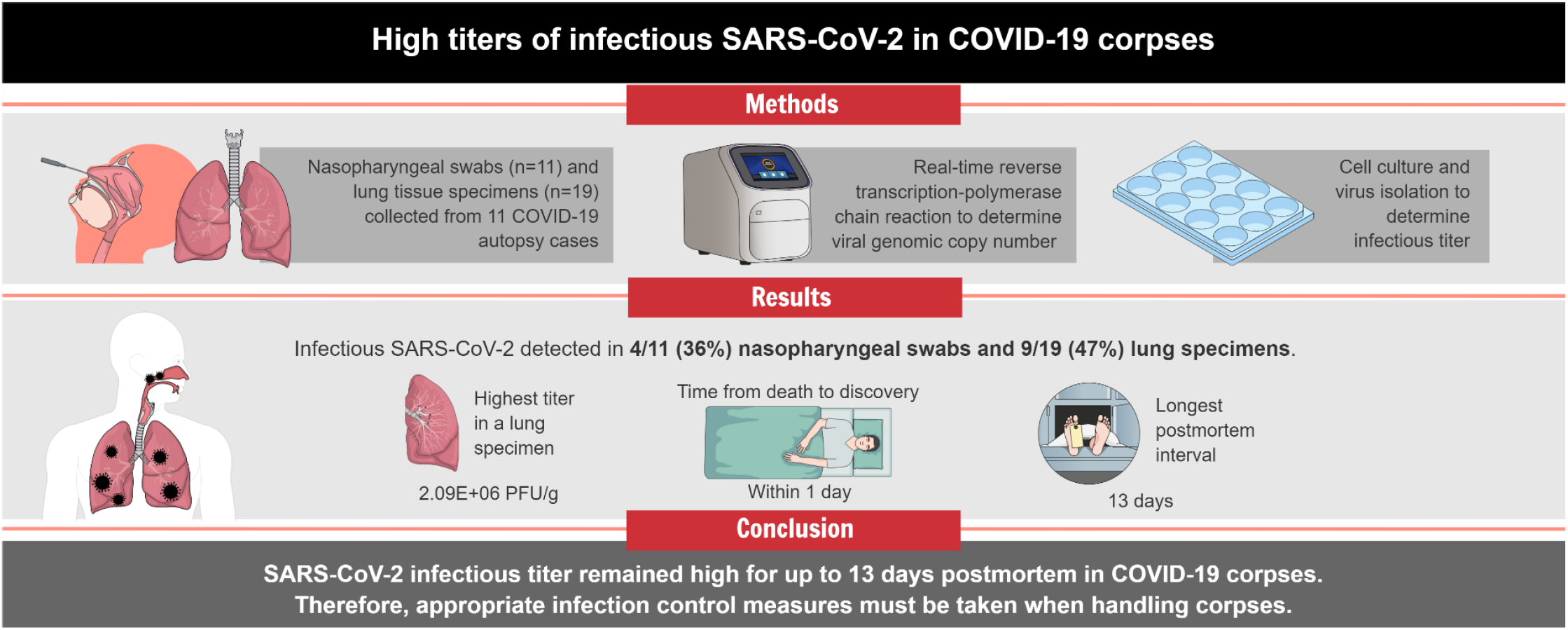

